# Intra-aortic balloon pump is associated with the lowest whereas Impella with the highest inpatient mortality and complications regardless of severity or hospital types (rural or university) or after adjustment for 47 high risk variables and baseline characteristics by studying over 800,000 inpatients with cardiogenic shock

**DOI:** 10.1101/2024.01.17.24301446

**Authors:** Mohammad Reza Movahed, Armin Talle, Mehrtash Hashemzadeh

## Abstract

**Background:** Impella and intra-aortic balloon pumps (IABP) are commonly utilized in patients with cardiogenic shock. However, the effect on mortality remains controversial. The goal of this study was to evaluate the effect of Impella and IABP on mortality and complications in patients with cardiogenic shock.

**Method:** The large Nationwide Inpatient Sample (NIS) database was utilized to study any association between the use of IABP or Impella on mortality and complications in adults with a diagnosis of cardiogenic shocks.

**Results:** ICD-10 codes for Impella, IABP, and cardiogenic shock for available years 2016-2020 were utilized. A total of 844,020 patients had a diagnosis of cardiogenic shock. 101,870 were treated with IABP and 39,645 with an Impella. Total inpatient mortality without any device was 34.2% vs only 25.1% with IABP use (OR=0.65, CI 0.62-0.67) but was highest at 40.7% with Impella utilization (OR=1.32, CI 1.26-1.39). Using multivariate analysis adjusting for 47 variables such as age, gender, race, lactose acidosis, three-vessel intervention, left main myocardial infarction, cardiomyopathy, systolic heart failure, acute ST-elevation myocardial infarction, peripheral vascular disease, chronic renal disease, etc., Impella utilization remained associated with the highest mortality (OR: 1.33, CI 1.25-1.41, p<0.001) whereas IABP remained associated with the lowest mortality (OR: 0.69, CI 0.66-0.72, p<0.001). Separating rural vs teaching hospitals revealed similar findings.

**Conclusion:** In patients with cardiogenic shock, the use of Impella was associated with the highest whereas the utilization of IABP was associated with the lowest in hospital mortality regardless of comorbid condition, high-risk futures, or type of hospital.

## Introduction

Cardiogenic shock (CS) is defined by SBP <90 mm Hg for 30 min or inotropes use to maintain SBP >90 mm Hg and evidence of end□organ damage and increased filling pressures a life-threatening condition characterized by severe cardiac dysfunction leading to inadequate tissue perfusion. It occurs in 5-15% of patients following a myocardial infarction (MI) and remains the leading cause of mortality^1^. The management of cardiogenic shock often involves the use of mechanical circulatory support (MCS) devices to augment cardiac function and improve hemodynamic stability^2^. Among the commonly utilized devices are the intra-aortic balloon pump (IABP) and the Impella (Extracorporeal membrane oxygenation, ECMO is also available but less commonly utilized due to availability and complexity of use). These devices aim to provide temporary circulatory support and enhance cardiac output in patients with compromised cardiac function. However, the impact of IABP and Impella utilization on mortality in patients with cardiogenic shock remains an area of ongoing investigation with controversial reports with no clear answer regarding improvement in outcome.

The IABP introduced nearly 5 decades ago, is the most widely used MCS device. Despite this widespread use, supportive evidence of significant improvement in mortality in these patients has been limited. In fact, the IABP Shock II Trial (n=600), the first RCT on the efficacy of IABP, revealed no significant improvement in 30-day mortality in patients with cardiogenic shock complicating acute MI^2,3^. This trial had a recent 6-year follow-up study completed which also showed significant differences in mortality, recurrent MI, stroke, repeat revascularization, or rehospitalization for cardiac reasons^4^. However, this was a small study with many patients enrolled who had a low-risk or borderline cardiogenic shock. While more recent data has been published demonstrating the efficacy of IABPs in the reduction of 30-day and 1-year mortality in CS patients, the inconsistency of the data and significance of the IABP Shock II follow-up has led to European Society of Cardiology (ESC) and European Association for Cardio-Thoracic Surgery (EACTS) guidelines downgrading the use of these devices from class I to a class III B recommendation (Not Recommended for routine use in cardiogenic shock due to ACS) and American College of Cardiology Foundation (ACCF) and the American Heart Association (AHA) guidelines downgrading it to a class IIb B recommendation (Weak/Usefulness is unknown/unclear/uncertain)^5–8^ This downgrading appears to be not justified as the largest meta-analysis involving 10,985 patients with cardiogenic shock comparing IABP with ECMO and Impella, IABP was found to be superior in improving mortality compared to Impella and ECMO. ^9^ Data from the NIS database from 2005-2014 (n= 144,254) revealed a significant downward trend of IABP use in CS patients (29.8–17.7%) and a significant uptrend in Impella (0.1–2.6%) and ECMO (0.3–1.8%) usage^10^. The first Impella model received FDA approval for use in the US in 2008. These devices are axial flow pumps that are advanced from the common femoral artery and pass retrograde across the aortic valve into the LV and eject blood into the ascending aorta. While the vast majority of data regarding its efficacy in the treatment of CS has been observational and registry data, the promising data has led to widespread use in 1000+ facilities countrywide^10–13^. Despite the trends in utilization, there remains a need for further investigation on the effectiveness of Impella vs IABP in the treatment of cardiogenic shock. The current literature on this comparison began with the IMPRESS trial (n=46) in 2016 which showed all-cause mortality for Impella CP vs. IABP was 46% vs. 50%^14^. Subsequent larger trials range from no significant improvement with either method, improvement in IABP compared to Impella, and increased risk of adverse events such as bleeding, sepsis, or limb complications^14–17^.

However, even the largest of these trials only includes 6885 Impella cases. Additionally, recent randomized trials and retrospective systemic reviews have suggested both patient harm and increased hospital costs with the use of Impella devices^19^. Major complications that were shown in these various studies included increased in-hospital mortality, major bleeding and the need for transfusions, limb complications, and hemolysis^14–22^

Therefore, a comprehensive evaluation of a large patient population is essential to provide more definitive evidence on the comparative mortality outcomes associated with IABP and Impella utilization. The objective of this study was to evaluate the association between the utilization of IABP or Impella and mortality in a large cohort of patients with cardiogenic shock. To accomplish this, we retrospectively analyzed data from the Nationwide Inpatient Sample (NIS) database of adult patients to achieve the largest sample size in a study of this kind. By examining the inpatient mortality of patients treated with IABP or Impella, we aimed to shed light on the relative effectiveness of these devices in reducing mortality in cardiogenic shock. The findings of this study have the potential to inform clinical decision-making, optimize patient management, and improve overall outcomes in the treatment of cardiogenic shock.

## Methods

### Data source

The Nationwide Inpatient Sample (NIS) database used in this study was developed by the Agency for Healthcare Research and Quality (AHRQ) as part of the Healthcare Cost and Utilization Project (HCUP). The database is de-identified, exempt from IRB approval, and publicly available to researchers and policymakers for analysis of nationwide trends in healthcare utilization and outcomes. The NIS contains information on primary and secondary diagnoses and procedures, discharge vital status, and demographics from nearly 1/5th of all US community hospitals.

### Sample Selection

This retrospective study included patients age>18 discharged from a NIS hospital in 2016-2020 with the following International Classification of Diseases, Tenth Revision, and Clinical Modification (ICD-10-CM) codes: Cardiogenic Shock (R57.0), Balloon Pump (5A02210), Impella Pump (5A0221D). We performed multivariate analysis adjusting for 47 factors such as age, gender and race, and co-morbidities in order to exclude the effect of confounding factors. These comorbidities were as follows: smoking, diabetes mellitus (250), chronic kidney disease, Peripheral Vascular Diseases, Cardiomyopathy, Systolic Heart Failure, PCI Three Vessel, Left main STEMI, STEMI, Anterior Wall STEMI, Cachexia, Morbid Obesity, Obesity, Chronic Liver Disease, Atrial Fibrillation/Flutter, COPD, ALL Valvular Heart Disease, History of Stroke, Acute Lactic Acidosis, Cardiac Arrest, Mechanical Ventilation, Renal Replacement Therapy, Heart Failure, Presence of Aortocoronary Bypass Graft, Right Ventricular Infarction, Rotational Atherectomy (Table 1). Comorbidities with significant p values were put in multivariate analysis for further adjustment. Furthermore, we evaluated outcomes data in teaching hospitals vs rural.

**Table 1:**
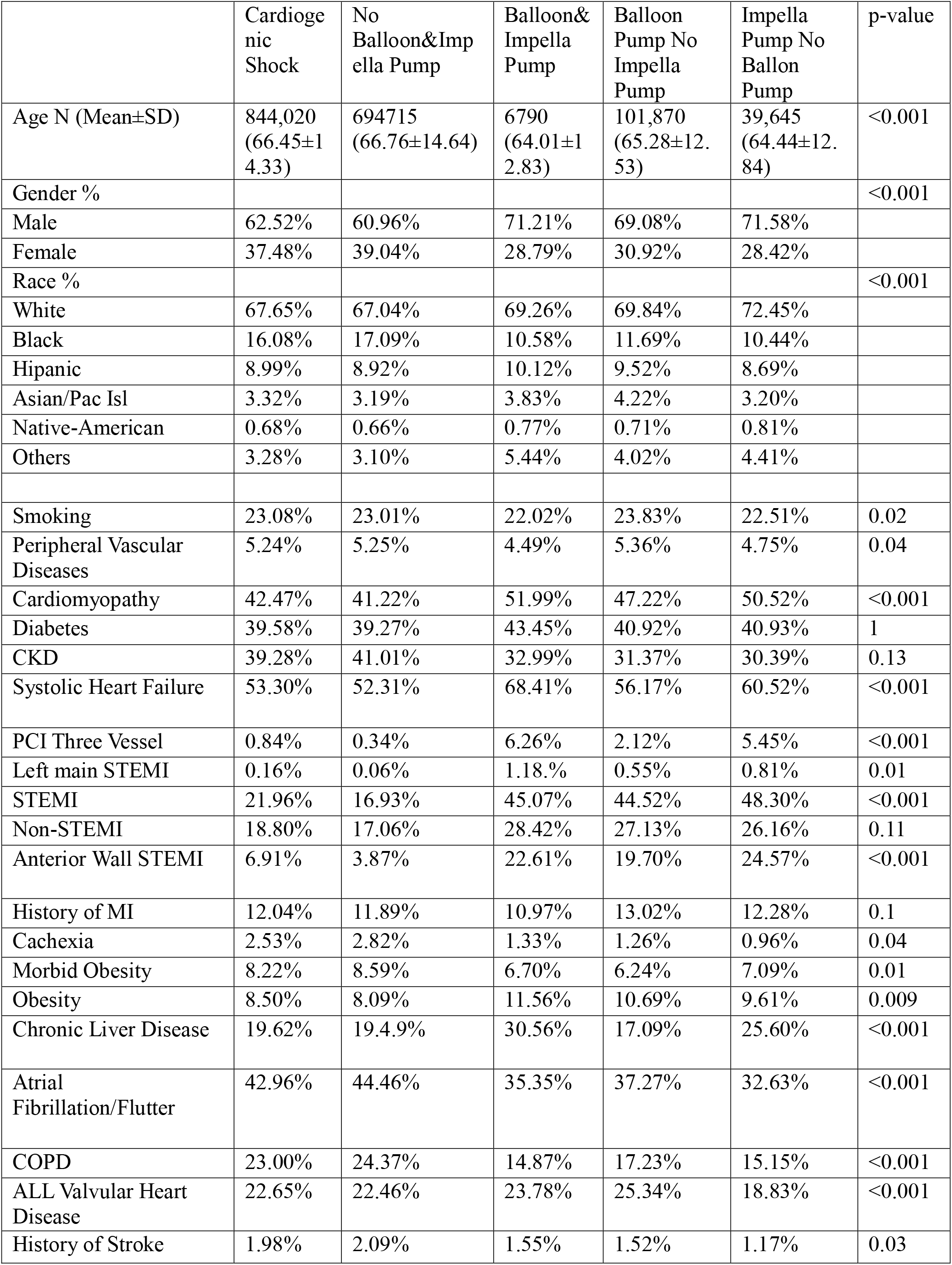
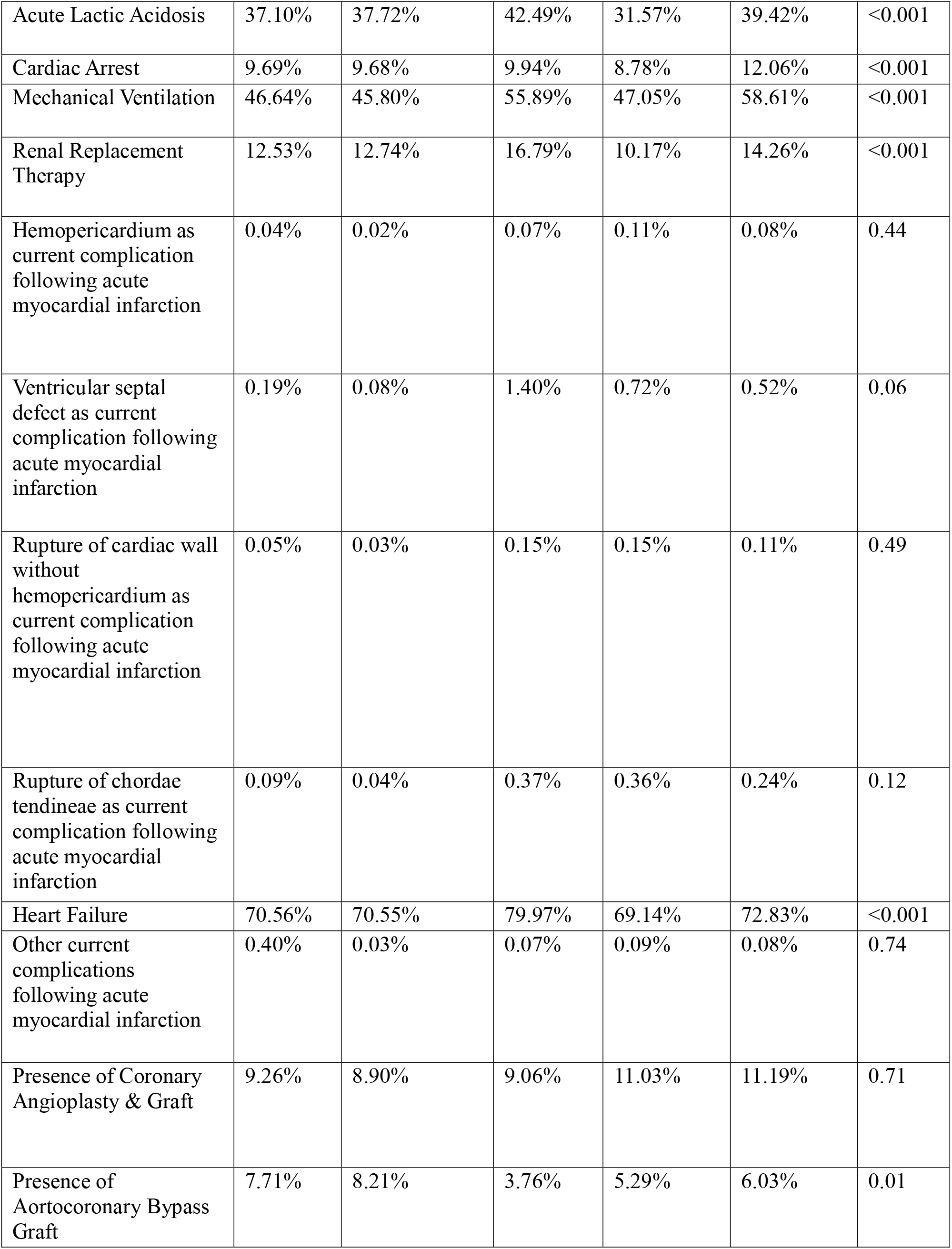
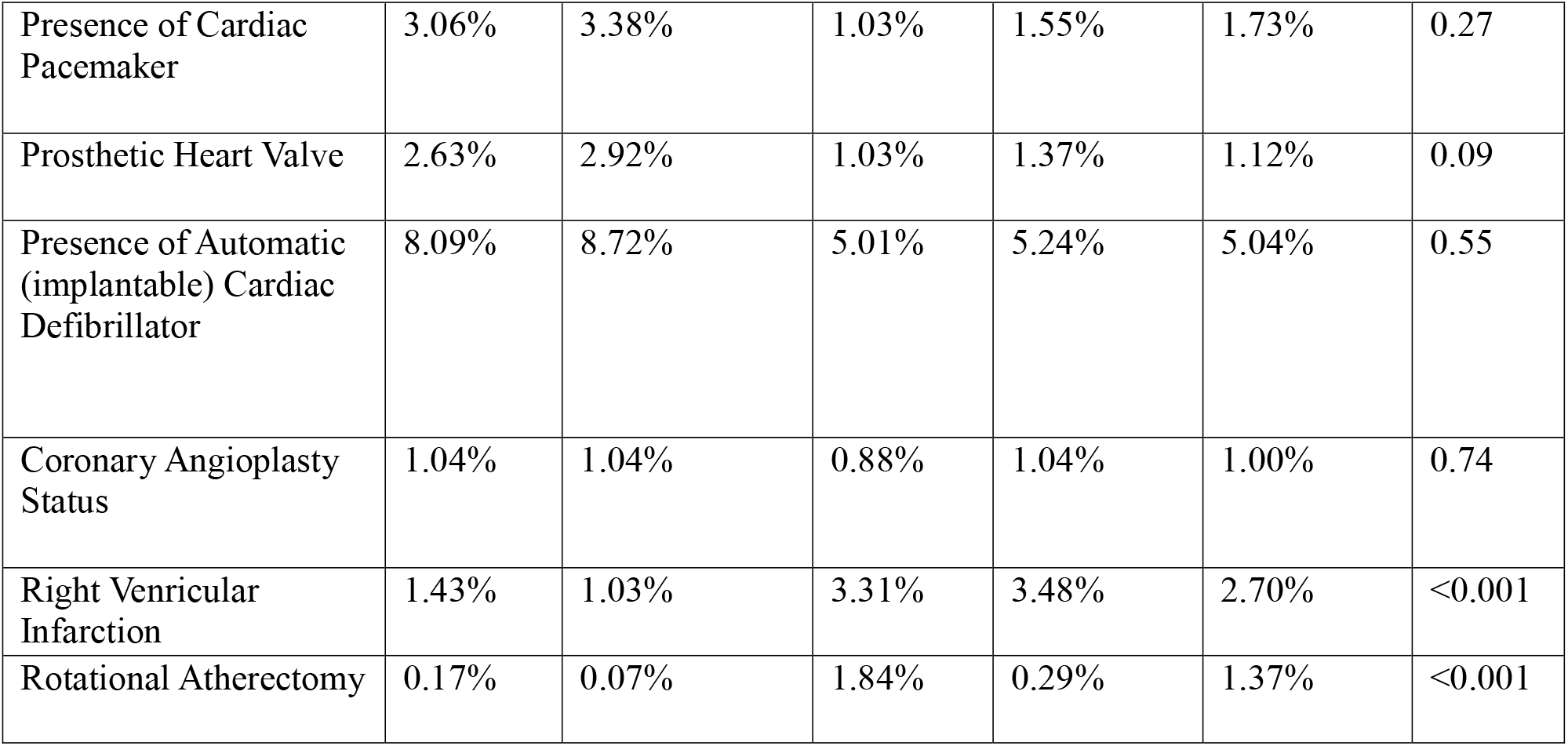
Baseline characteristic of patients with cardiogenic shock.

|  | Cardiogenic Shock | No Balloon&Impella Pump | Balloon&Impella Pump | Balloon Pump No Impella Pump | Impella Pump No Balloon Pump | p-value |
| --- | --- | --- | --- | --- | --- | --- |
| Age N (Mean±SD) | 844,020<br>(66.45±14.33) | 694715<br>(66.76±14.64) | 6790<br>(64.01±12.83) | 101,870<br>(65.28±12.53) | 39,645<br>(64.44±12.84) | <0.001 |
| Gender % |  |  |  |  |  | <0.001 |
| Male | 62.52% | 60.96% | 71.21% | 69.08% | 71.58% |  |
| Female | 37.48% | 39.04% | 28.79% | 30.92% | 28.42% |  |
| Race % |  |  |  |  |  | <0.001 |
| White | 67.65% | 67.04% | 69.26% | 69.84% | 72.45% |  |
| Black | 16.08% | 17.09% | 10.58% | 11.69% | 10.44% |  |
| Hipanic | 8.99% | 8.92% | 10.12% | 9.52% | 8.69% |  |
| Asian/Pac Isl | 3.32% | 3.19% | 3.83% | 4.22% | 3.20% |  |
| Native-American | 0.68% | 0.66% | 0.77% | 0.71% | 0.81% |  |
| Others | 3.28% | 3.10% | 5.44% | 4.02% | 4.41% |  |
| Smoking | 23.08% | 23.01% | 22.02% | 23.83% | 22.51% | 0.02 |
| Peripheral Vascular Diseases | 5.24% | 5.25% | 4.49% | 5.36% | 4.75% | 0.04 |
| Cardiomyopathy | 42.47% | 41.22% | 51.99% | 47.22% | 50.52% | <0.001 |
| Diabetes | 39.58% | 39.27% | 43.45% | 40.92% | 40.93% | 1 |
| CKD | 39.28% | 41.01% | 32.99% | 31.37% | 30.39% | 0.13 |
| Systolic Heart Failure | 53.30% | 52.31% | 68.41% | 56.17% | 60.52% | <0.001 |
| PCI Three Vessel | 0.84% | 0.34% | 6.26% | 2.12% | 5.45% | <0.001 |
| Left main STEMI | 0.16% | 0.06% | 1.18% | 0.55% | 0.81% | 0.01 |
| STEMI | 21.96% | 16.93% | 45.07% | 44.52% | 48.30% | <0.001 |
| Non-STEMI | 18.80% | 17.06% | 28.42% | 27.13% | 26.16% | 0.11 |
| Anterior Wall STEMI | 6.91% | 3.87% | 22.61% | 19.70% | 24.57% | <0.001 |
| History of MI | 12.04% | 11.89% | 10.97% | 13.02% | 12.28% | 0.1 |
| Cachexia | 2.53% | 2.82% | 1.33% | 1.26% | 0.96% | 0.04 |
| Morbid Obesity | 8.22% | 8.59% | 6.70% | 6.24% | 7.09% | 0.01 |
| Obesity | 8.50% | 8.09% | 11.56% | 10.69% | 9.61% | 0.009 |
| Chronic Liver Disease | 19.62% | 19.4.9% | 30.56% | 17.09% | 25.60% | <0.001 |
| Atrial Fibrillation/Flutter | 42.96% | 44.46% | 35.35% | 37.27% | 32.63% | <0.001 |
| COPD | 23.00% | 24.37% | 14.87% | 17.23% | 15.15% | <0.001 |
| ALL Valvular Heart Disease | 22.65% | 22.46% | 23.78% | 25.34% | 18.83% | <0.001 |
| History of Stroke | 1.98% | 2.09% | 1.55% | 1.52% | 1.17% | 0.03 |
| Acute Lactic Acidosis | 37.10% | 37.72% | 42.49% | 31.57% | 39.42% | <0.001 |
| Cardiac Arrest | 9.69% | 9.68% | 9.94% | 8.78% | 12.06% | <0.001 |
| Mechanical Ventilation | 46.64% | 45.80% | 55.89% | 47.05% | 58.61% | <0.001 |
| Renal Replacement Therapy | 12.53% | 12.74% | 16.79% | 10.17% | 14.26% | <0.001 |
| Hemopericardium as current complication following acute myocardial infarction | 0.04% | 0.02% | 0.07% | 0.11% | 0.08% | 0.44 |
| Ventricular septal defect as current complication following acute myocardial infarction | 0.19% | 0.08% | 1.40% | 0.72% | 0.52% | 0.06 |
| Rupture of cardiac wall without hemopericardium as current complication following acute myocardial infarction | 0.05% | 0.03% | 0.15% | 0.15% | 0.11% | 0.49 |
| Rupture of chordae tendineae as current complication following acute myocardial infarction | 0.09% | 0.04% | 0.37% | 0.36% | 0.24% | 0.12 |
| Heart Failure | 70.56% | 70.55% | 79.97% | 69.14% | 72.83% | <0.001 |
| Other current complications following acute myocardial infarction | 0.40% | 0.03% | 0.07% | 0.09% | 0.08% | 0.74 |
| Presence of Coronary Angioplasty & Graft | 9.26% | 8.90% | 9.06% | 11.03% | 11.19% | 0.71 |
| Presence of Aortocoronary Bypass Graft | 7.71% | 8.21% | 3.76% | 5.29% | 6.03% | 0.01 |
| Presence of Cardiac Pacemaker | 3.06% | 3.38% | 1.03% | 1.55% | 1.73% | 0.27 |
| Prosthetic Heart Valve | 2.63% | 2.92% | 1.03% | 1.37% | 1.12% | 0.09 |
| Presence of Automatic (implantable) Cardiac Defibrillator | 8.09% | 8.72% | 5.01% | 5.24% | 5.04% | 0.55 |
| Coronary Angioplasty Status | 1.04% | 1.04% | 0.88% | 1.04% | 1.00% | 0.74 |
| Right Ventricular Infarction | 1.43% | 1.03% | 3.31% | 3.48% | 2.70% | <0.001 |
| Rotational Atherectomy | 0.17% | 0.07% | 1.84% | 0.29% | 1.37% | <0.001 |

### Statistical Analysis

Patient demographic, clinical, and hospital characteristics are reported as percentages in the tables. Odds ratios and 95% confidence intervals are calculated for continuous variables and proportions, and 95% confidence intervals for categorical variables. Trend analysis over time was assessed using Chi-squared analysis for categorical outcomes and univariate linear regression for continuous variables. Multivariable logistic regression ascertained the odds of binary clinical outcomes relative to patient and hospital characteristics as well as the odds of clinical outcomes over time. All analyses were conducted following the implementation of population discharge weights. All p-values are 2-sided and p<0.05 was considered statistically significant. Data were analyzed using STATA 17 (Stata Corporation, College Station, TX). The occurrence of cardiogenic shock and in-hospital mortality rates were calculated for each year for trends (2016-2020) and all together for final analysis.

## Results

### Mortality

A total of 844,020 patients had a diagnosis of cardiogenic shock from 2016-2020. Complete demographic data are shown in Table 1. Of the sample, 101,870 (mean age 65.2) were treated with IABP, and 39,645 (mean age 64.4) were treated with an Impella. Overall inpatient mortality was 33.54%. Inpatient mortality in patients without mechanical circulatory support was 34.22%. Inpatient mortality in IABP alone was markedly lower at 25.19% (p<0.001, OR: 0.65(0.62-0.67)) whereas inpatient mortality in Impella was markedly higher at 40.75% (p<0.001 OR: 1.32(1.26-1.39)). Inpatient mortality in Impella plus IABP was the highest at 46.83% (p<0.001 OR: 2.12 (1.89-2.37)). The mortality trends over the years remained stable. Yearly trends are seen in Figure 1. We hypothesized that due to the lack of Impella availability in rural hospital (rural hospitals used Impella only in 3.4% of cardiogenic shock patients), many high-risk patients will undergo IABP insertion and therefore, mortality of IABP in this population should be higher than no device. However, we find the same consistent finding that IABP reduced mortality similar to the entire cohort (IABP mortality 25.18% vs 36.01% on no device OR 0.60 CI:0.50-0.71, P<0.001) Furthermore, we assumed that the university hospitals should provide better care in patients with Impella insertion that could lead to lower mortality in this patient. However, the results of Impella use in the teaching hospital was also similar to the entire cohort increasing mortality by approximately 40% vs no device use (Impella mortality 41.12% vs 33.60% with no device OR 1.38, CI:1.31-1.46, p <000.1)

**Figure 1:**
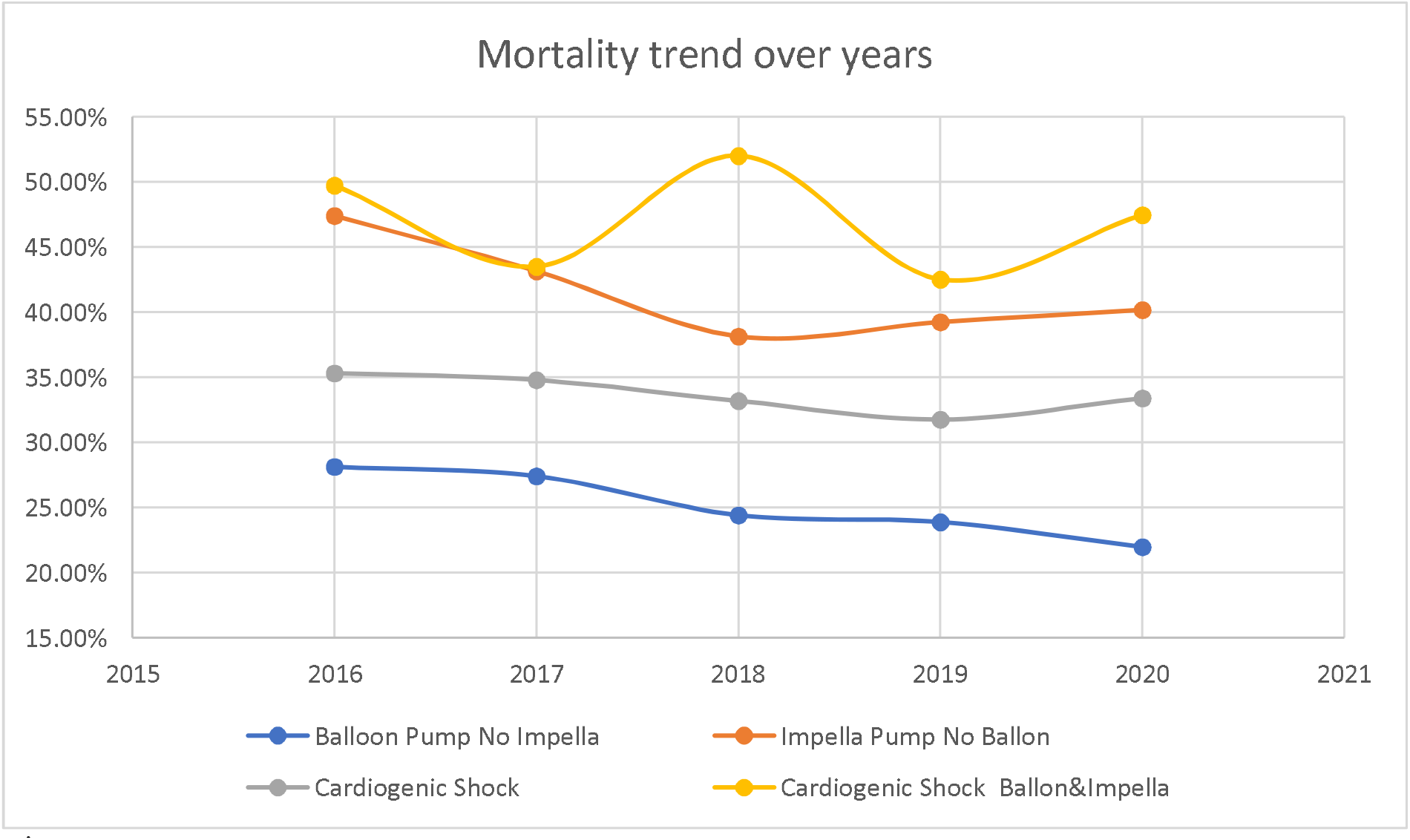

### Multivariate and Subgroup Analysis

Using multivariate analysis adjusting for multiple demographics and patient-specific factors, Impella utilization remained associated with the highest mortality (OR: 1.33, CI 1.25-1.41, p<0.001) whereas IABP remained associated with the lowest mortality (OR: 0.69, CI 0.66-0.72, p<0.001). In order to study any subgroups that may benefit from Impella or have harm done by IABP insertion, we evaluated mortality in different demographics and in high-risk patients. As can be seen in Figure 2 and Figure 3, Impella significantly increased mortality in all subgroups whereas IABP used reduced mortality in all subgroups studied.

**Figure 2:**
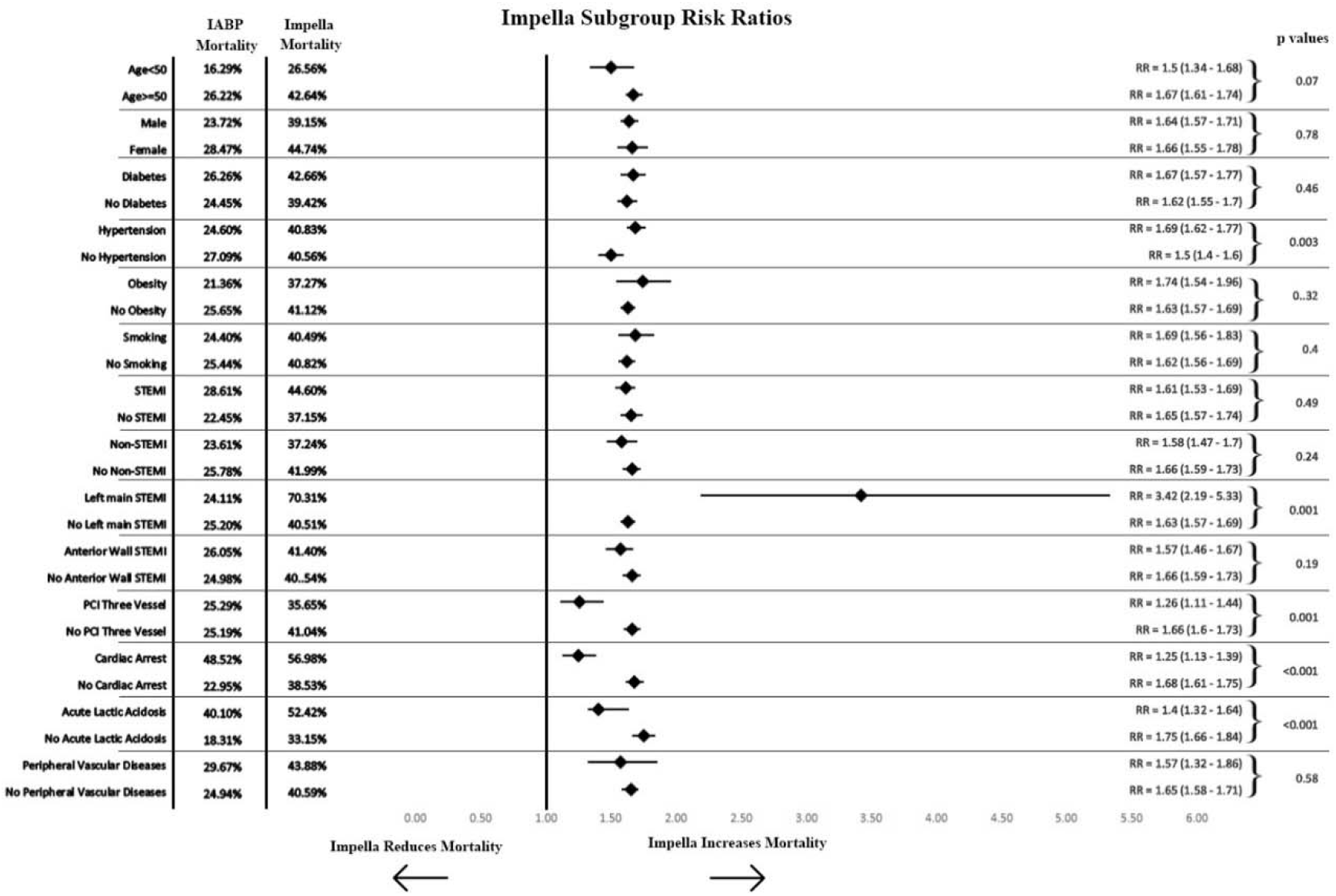
Mortality is higher with the use of Impella regardless of any subgroup.

**Figure 3:**
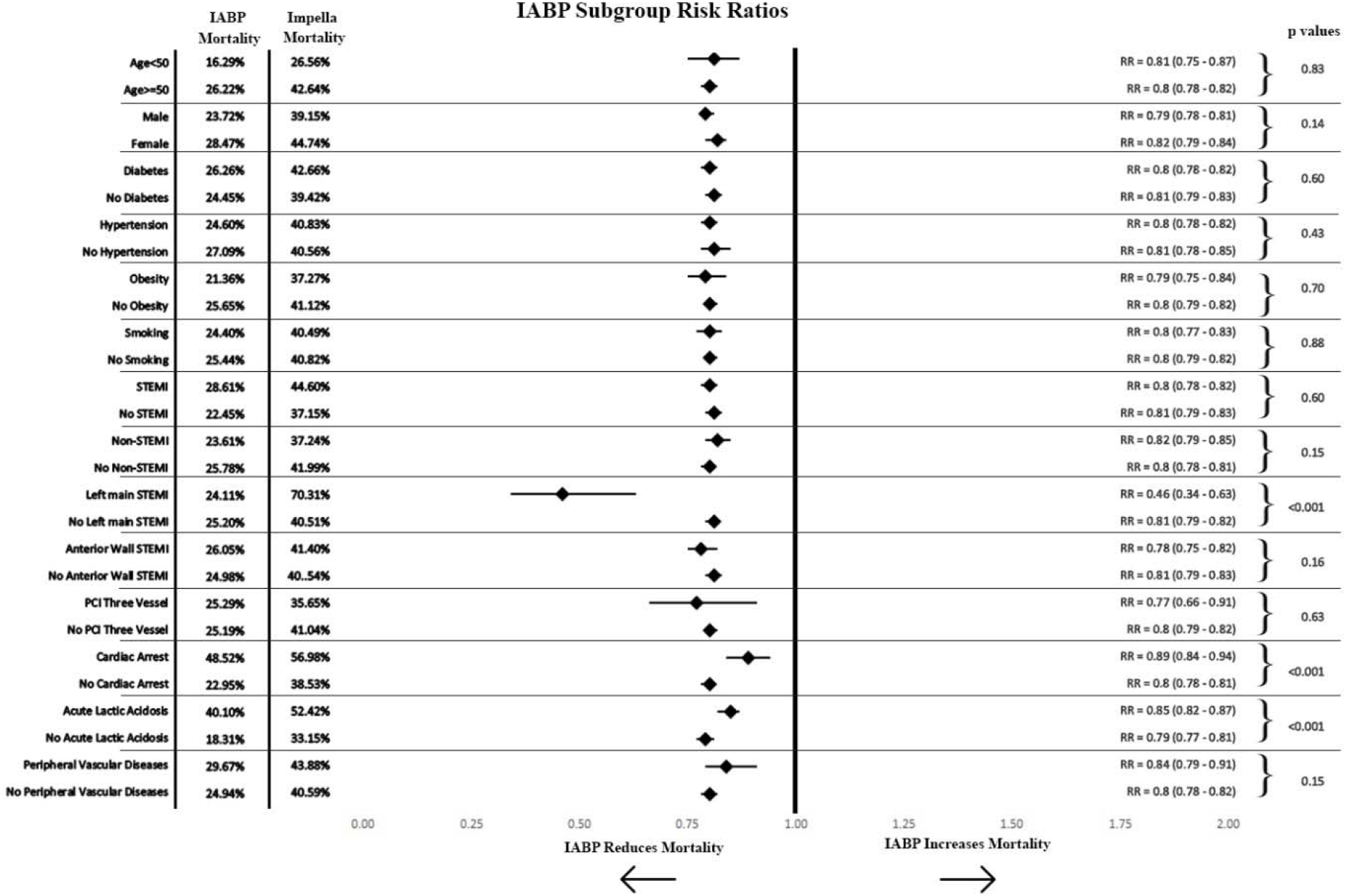
Mortality is lower with the use of IABP regardless of any subgroup.

### Complications

Patients who received Impella had a significant increase in various complications compared to IABP including Cardiac Tamponade (OR= 1.27(1.07-1.51), p=0.007), Hemolytic anemia (OR= 8.36(4.89-14.30), p <0.001), Post-procedural hemorrhage (OR= 1.99(10.64-2.42), p <0.001), Disseminated Intravascular Coagulation (OR= 1.76(1.48-2.08, p <0.001), Cardiac perforation (OR= 1.48(1.17-1.88), p<0.001) and Procedural bleeding (OR= 2.37(1.33-4.23), p=0.004) in comparison to IABP use. Patients who received IABP had significant increases in Post-procedural AKI and Acute post-procedural respiratory failure in comparison to Impella(Table 2).

**Table 2:**
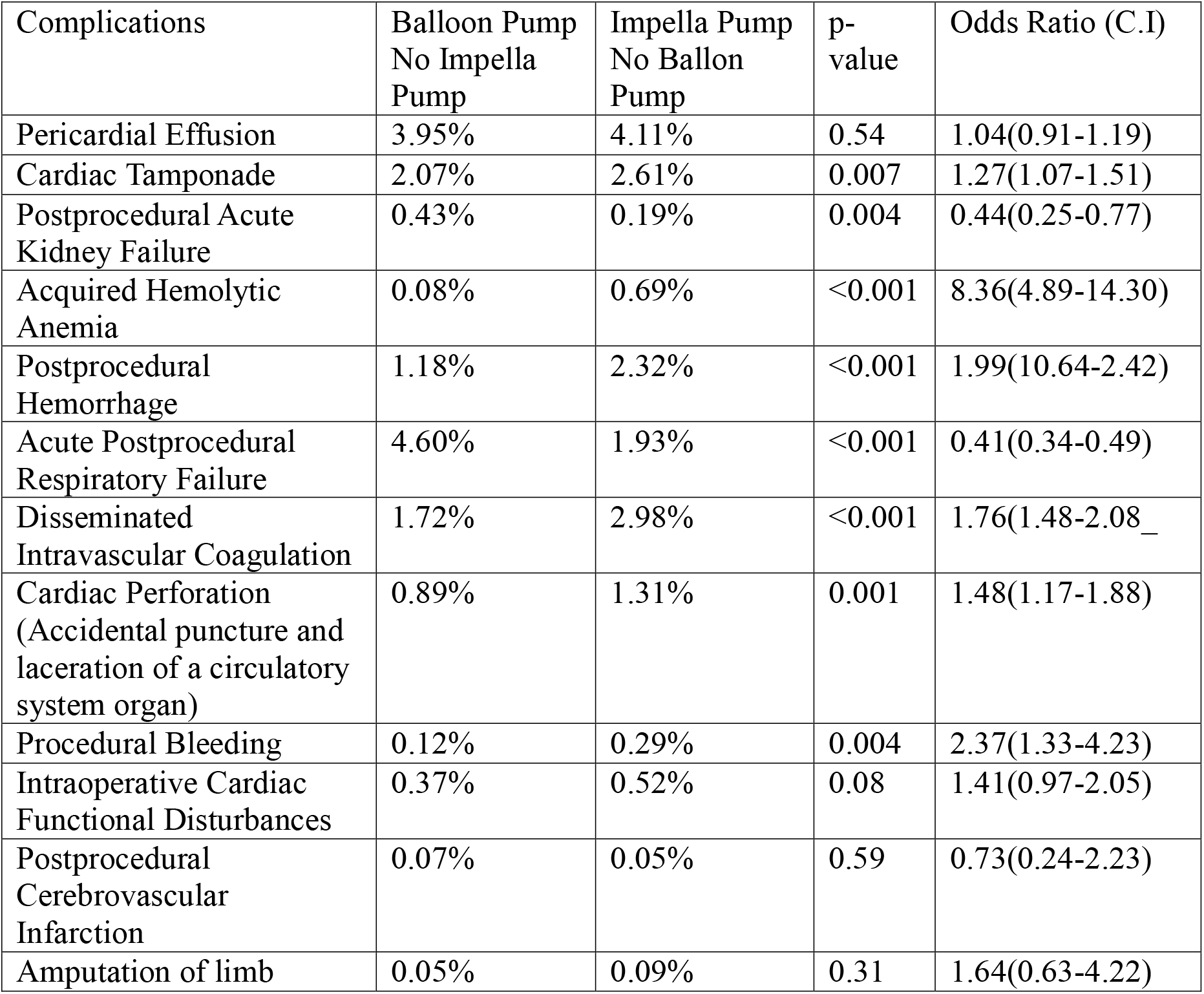
Complications in patients with Impella vs intra-aortic balloon-pump insertion.

## Discussion

The management of cardiogenic shock often involves the use of mechanical circulatory support (MCS) devices to augment cardiac function and improve hemodynamic stability. The IABP and Impella are two commonly utilized devices for this purpose. However, the impact of these devices on mortality in patients with cardiogenic shock has been a subject of ongoing investigation, with conflicting reports and no clear consensus on their effectiveness. The use of Impella in this patient population continues to gain popularity over the Intra-Aortic Balloon Pump^9,21^. While randomized control trials and large-scale retrospective analysis have shown demonstrated its effectiveness in high-risk PCI cases, the data on treatment for cardiogenic shock has not been well-established^22-24^. The downgrading of IABP use has led to a steady decrease in the utilization of these devices in patients with cardiogenic shock. However, significant weight was placed on the SHOCK II randomized control trial consisting only of 600 patients with many having low-risk cardiogenic shock features. ^4,25^. We present the largest retrospective review of the use of IABP and Impella to provide valuable insight into the efficacy of these modalities.

Our study found that in patients with cardiogenic shock, the use of Impella was associated with significant increases in in-hospital mortality compared to no treatment. Additionally, the use of IABP was associated with a significant decrease in in-hospital mortality. This trend was even more pronounced when looking at the cardiogenic shock in the high-risk subgroup Left Main STEMI vs No Left Main STEMI with IABP further significantly improving mortality and Impella further significantly increased mortality in these cases. Consistent with much of the current literature on this patient population, Impella use was associated with significantly more life-threatening conditions compared to IABP use^15–22^. Impella was found to be associated with an 8-fold increase in the prevalence of hemolytic anemia, as well as a 2-fold increase in procedural and postprocedural bleeding and DIC. These findings potentially explain the observed trends in mortality. Impella device is a bulky device that utilizes large access that can increase the risk of severe bleeding and leg ischemia. It also requires significant anticoagulation whereas anticoagulation in patients with IABP is optional. Furthermore, Impella requires insertion of the device across the aortic valve which can increase myocardial damage or arrhythmia. The fact that we found higher perforations in the Impella group suggests that perforation may play an important role in observed increased mortality.

Also, there is some evidence that the use of stronger Impella devices such as the Impella 5.0 and Impella CP may improve clinical outcomes and our database is unable to divide the Impella group into specific types^26^. However, we utilized extensive multivariate analysis to control for 47 possible confounding variables including markers for high-risk factors and still saw consistent significant results that point to increased mortality in Impella cases regardless of subgroup division and improved mortality in IABP cases in every subgroup. Our data suggest that the observed trends in mortality associated with IABP and Impella in patients with cardiogenic shock are independent of patient selection. This argues against the recent downgrading of IABP and suggests we could both improve patient outcomes and reduce unnecessary costs through the use of IABP for patients in cardiogenic shock.

There could be an argument that we could not possibly adjust for many unknown factors and higher mortality in the Impella arms could be related to much sicker patients leading to Impella insertion. It is true that we cannot rule out the fact that Impella patients could have been much sicker which may explain higher mortality but this fact should be also valid for patients undergoing IABP insertion. However, if patients with IABP use were much sicker than expected and IABP should be useless, then in comparison to no device use, IABP should have shown much higher mortality. The fact that the use of IABP was associated with much lower mortality despite the fact that much sicker patients should undergo IABP insertion in comparison to no device use suggests that the severity of cardiogenic shock is less likely the explanation of IABP benefit and higher mortality in Impella patients. For the same reason, we evaluated the effect of IABP in rural hospitals where most hospitals do not have Impella availability and assumed that much sicker patients undergoing IABP should have higher mortality but we saw a similar dramatic reduction in mortality of IABP in rural hospitals with an OR of 0.6. Furthermore, we evaluated Impella use in teaching hospitals with the assumption that they have a much better experience that can lead to lower mortality in the Impella arm. However, our results showed that higher mortality remains consistent even in teaching hospitals suggesting higher mortality seen with Impella use is most likely related to the Impella itself. In order to have a definitive answer, large randomized trials are needed to evaluate the effect of IABP and Impella on mortality in patients with cardiogenic shock. Our results are consistent with the largest metanalysis of 10,985 patients with cardiogenic shock showing superiority of IABP regarding to mortality compered to ECMO and Impella. ^9^ A recently published randomized study using Extracorporeal Life Support in Infarct-Related Cardiogenic revealed higher complications with this bulky device similar to Impella with no mortality benefit consistent with our finding. ^27^

### Limitations

We used ICD-10 coding which has inherent limitations in accurate diagnosis. Furthermore, we cannot exclude that patients who underwent Impella insertion were much sicker which we could not capture in our 47 variables. However, as mentioned above, the fact that patients with IABP insertion had much lower mortality despite the same fact that they should have been a much sicker population suggests that our data are valid. Furthermore, can not assess how clinicians came to their decision on whether to use either an Impella, IABP, or no device in each patient.

## Conclusion

In patients with cardiogenic shock, the use of Impella was associated with the highest inpatients mortality whereas the utilization of IABP was associated with the lowest mortality regardless of comorbid condition or any high-risk subgroup analysis. The largest randomized trial is required to definitely answer the effect of IABP or Impella in patients with cardiogenic shocks.

## Data Availability

Data publically available NIS database

## References

1. Elgendy IY, Van Spall HGC, Mamas MA. Cardiogenic Shock in the Setting of Acute Myocardial Infarction. Circ Cardiovasc Interv. 2020;13(3):e009034. doi:10.1161/CIRCINTERVENTIONS.120.009034

2. Vahdatpour C, Collins D, Goldberg S. Cardiogenic Shock. J Am Heart Assoc. 2019;8(8):e011991. doi:10.1161/JAHA.119.011991

3. Thiele H, Zeymer U, Neumann FJ, et al. Intraaortic Balloon Support for Myocardial Infarction with Cardiogenic Shock. N Engl J Med. 2012;367(14):1287–1296. doi:10.1056/NEJMoa1208410

4. Thiele H, Zeymer U, Thelemann N, et al. Intraaortic Balloon Pump in Cardiogenic Shock Complicating Acute Myocardial Infarction. Circulation. 2019;139(3):395–403. doi:10.1161/CIRCULATIONAHA.118.038201

5. Zein R, Patel C, Mercado-Alamo A, Schreiber T, Kaki A. A Review of the Impella Devices. Interv Cardiol Rev Res Resour. 2022;17:e05. doi:10.15420/icr.2021.11

6. Yuan S, He J, Cai Z, et al. Intra-aortic balloon pump in cardiogenic shock: A propensity score matching analysis. Catheter Cardiovasc Interv Off J Soc Card Angiogr Interv. 2022;99 Suppl 1:1456–1464. doi:10.1002/ccd.30102

7. Neumann FJ, Sousa-Uva M, Ahlsson A, et al. 2018 ESC/EACTS Guidelines on myocardial revascularization. Eur Heart J. 2019;40(2):87–165. doi:10.1093/eurheartj/ehy394

8. Tehrani BN, Truesdell AG, Psotka MA, et al. A Standardized and Comprehensive Approach to the Management of Cardiogenic Shock. JACC Heart Fail. 2020;8(11):879–891. doi:10.1016/j.jchf.2020.09.005

9. Zhang Q, Han Y, Sun S, Zhang C, Liu H, Wang B, Wei S. Mortality in cardiogenic shock patients receiving mechanical circulatory support: a network meta-analysis. BMC Cardiovasc Disord. 2022 Feb 13;22(1):48

10. Shah M, Patnaik S, Patel B, et al. Trends in mechanical circulatory support use and hospital mortality among patients with acute myocardial infarction and non-infarction related cardiogenic shock in the United States. Clin Res Cardiol. 2018;107(4):287–303. doi:10.1007/s00392-017-1182-2

11. Glazier JJ, Kaki A. The Impella Device: Historical Background, Clinical Applications and Future Directions. Int J Angiol Off Publ Int Coll Angiol Inc. 2019;28(2):118–123. doi:10.1055/s-0038-1676369

12. Lemaire A, Anderson MB, Lee LY, et al. The Impella Device for Acute Mechanical Circulatory Support in Patients in Cardiogenic Shock. Ann Thorac Surg. 2014;97(1):133–138. doi:10.1016/j.athoracsur.2013.07.053

13. O’Neill WW, Schreiber T, Wohns DHW, et al. The current use of Impella 2.5 in acute myocardial infarction complicated by cardiogenic shock: results from the USpella Registry. J Intervent Cardiol. 2014;27(1):1–11. doi:10.1111/joic.12080

14. Ouweneel DM, Eriksen E, Sjauw KD, et al. Percutaneous Mechanical Circulatory Support Versus Intra-Aortic Balloon Pump in Cardiogenic Shock After Acute Myocardial Infarction. J Am Coll Cardiol. 2017;69(3):278–287. doi:10.1016/j.jacc.2016.10.022

15. Moustafa A, Khan MS, Saad M, Siddiqui S, Eltahawy E. Impella Support Versus Intra-Aortic Balloon Pump in Acute Myocardial Infarction Complicated by Cardiogenic Shock: A Meta-Analysis. Cardiovasc Revascularization Med Mol Interv. 2022;34:25–31. doi:10.1016/j.carrev.2021.01.028

16. Frain K, Rees P. Intra-aortic balloon pump versus percutaneous Impella© in emergency revascularisation for myocardial infarction and cardiogenic shock: systematic review. Perfusion. Published online September 3, 2021:2676591211037026. doi:10.1177/02676591211037026

17. Alushi B, Douedari A, Froehlig G, et al. Impella versus IABP in acute myocardial infarction complicated by cardiogenic shock. Open Heart. 2019;6(1):e000987. doi:10.1136/openhrt-2018-000987

18. Jin C, Yandrapalli S, Yang Y, Liu B, Aronow WS, Naidu SS. A Comparison of In-Hospital Outcomes Between the Use of Impella and IABP in Acute Myocardial Infarction Cardiogenic Shock Undergoing Percutaneous Coronary Intervention. J Invasive Cardiol. 2022;34(2):E98–E103.

19. Kim Y, Shapero K, Ahn SS, Goldsweig AM, Desai N, Altin SE. Outcomes of mechanical circulatory support for acute myocardial infarction complicated by cardiogenic shock. Catheter Cardiovasc Interv Off J Soc Card Angiogr Interv. 2022;99(3):658–663. doi:10.1002/ccd.29834

20. Bochaton T, Huot L, Elbaz M, et al. Mechanical circulatory support with the Impella® LP5.0 pump and an intra-aortic balloon pump for cardiogenic shock in acute myocardial infarction: The IMPELLA-STIC randomized study. Arch Cardiovasc Dis. 2020;113(4):237–243. doi:10.1016/j.acvd.2019.10.005

21. Kuno T, Takagi H, Ando T, et al. Safety and efficacy of mechanical circulatory support with Impella or intra-aortic balloon pump for high-risk percutaneous coronary intervention and/or cardiogenic shock: Insights from a network meta-analysis of randomized trials. Catheter Cardiovasc Interv. 2021;97(5):E636–E645. doi:10.1002/ccd.29236

22. Philipson DJ, Cohen DJ, Fonarow GC, Ziaeian B. Analysis of Adverse Events Related to Impella® Usage (From the Manufacturer and User Facility Device Experience and National Inpatient Sample Databases). Am J Cardiol. 2021;140:91–94. doi:10.1016/j.amjcard.2020.10.056

23. O’Neill WW, Kleiman NS, Moses J, et al. A prospective, randomized clinical trial of hemodynamic support with Impella 2.5 versus intra-aortic balloon pump in patients undergoing high-risk percutaneous coronary intervention: the PROTECT II study. Circulation. 2012;126(14):1717–1727. doi:10.1161/CIRCULATIONAHA.112.098194

24. Lansky AJ, Tirziu D, Moses JW, et al. Impella Versus Intra-Aortic Balloon Pump for High-Risk PCI: A Propensity-Adjusted Large-Scale Claims Dataset Analysis. Am J Cardiol. 2022;185:29–36. doi:10.1016/j.amjcard.2022.08.032

25. Unverzagt S, Buerke M, Waha A de, et al. Intra□aortic balloon pump counterpulsation (IABP) for myocardial infarction complicated by cardiogenic shock. Cochrane Database Syst Rev. 2015;(3). doi:10.1002/14651858.CD007398.pub3

26. Panuccio G, Neri G, Macrì LM, Salerno N, De Rosa S, Torella D. Use of Impella device in cardiogenic shock and its clinical outcomes: A systematic review and meta-analysis. Int J Cardiol Heart Vasc. 2022;40:101007. doi:10.1016/j.ijcha.2022.101007

27. Thiele H, Zeymer U, Akin I, Behnes M, Rassaf T, Mahabadi AA, Lehmann R, Eitel I, Graf T, Seidler T, Schuster A, Skurk C, Duerschmied D, Clemmensen P, Hennersdorf M, Fichtlscherer S, Voigt I, Seyfarth M, John S, Ewen S, Linke A, Tigges E, Nordbeck P, Bruch L, Jung C, Franz J, Lauten P, Goslar T, Feistritzer HJ, Pöss J, Kirchhof E, Ouarrak T, Schneider S, Desch S, Freund A; ECLS-SHOCK Investigators. Extracorporeal Life Support in Infarct-Related Cardiogenic Shock. N Engl J Med. 2023 Aug 26. doi: 10.1056/NEJMoa2307227

